# Cognitive change over one year among older adults with HIV and a low nadir CD4 cell count

**DOI:** 10.1101/2023.05.18.23290138

**Authors:** Marie-Josée Brouillette, Laurence Forcellino, Sybil Goulet-Stock, Lesley K Fellows, Lisa Koski, Marina B. Klein, Nancy E. Mayo

## Abstract

**Background:** Evidence regarding the risk of cognitive decline conferred by a low nadir CD4 cell count and increasing age in people living with HIV is mixed. The objective of this study was to assess the change in cognition over one year among older adults with well-controlled HIV infection and a history of low nadir CD4 cell count compared with the change in a matched non-HIV sample.

**Methods:** We recruited 50 HIV+ aviremic individuals 40 years or older, on stable antiretroviral treatment and with a nadir CD4 < 200 cells/μL, and seventeen matched HIV-negative individuals. Neuropsychological testing was performed twice, one year apart; an NPZ was computed by averaging all z-scores and five existing algorithms for a diagnosis of HAND were applied. Change was defined as making a reliable change on the NPZ or a change in HAND category (impaired vs not).

**Results:** Change in NPZ over one year was more often in the direction of an improvement, and not different between HIV+ and HIV-individuals. Among the HIV+, the proportion meeting criteria for HAND at baseline ranged from 34-80% depending on the classification algorithm. A reliable change in NPZ was demonstrated in a single HIV+ participant. In contrast, a transition between HAND category at one year was common.

**Conclusion:** Among aviremic HIV+ older adults with a history of low nadir CD4 cell count, change in NPZ over 1 year was comparable to that seen among demographically matched HIV-individuals and did not represent a reliable change while transition across HAND category was common. Rates of HAND were very dependent on the classification algorithm applied. These findings provide some explanation for the inconsistent findings from existing studies and highlight the importance of exercising caution when pooling results in the field of neuroHIV.

## Background

While the introduction of combination antiretroviral therapy (cART) has been accompanied by a marked decrease in HIV-associated mortality and morbidity, HIV-related cognitive complications have persisted, with reported rates of HIV-Associated Neurocognitive Disorder (HAND) of 30-50% (1, 2). The presence of cognitive impairment, even mild, is associated with reduced occupational and social function, thus interfering with the attainment of the World Health Organization ‘4^th^ 90 goal’ of achieving good health-related quality of life in at least 90% of those living with HIV (3-10).

Existing evidence suggests that people living with HIV who are well-treated show little decline in neuropsychological (NP) performance over time (11-16), although longitudinal data are sparse. However, it is unclear if this favorable course extends to those who are older, a group that also typically acquired HIV prior to the cART era, and thus tends to have low nadir CD4 cell counts. Evidence regarding the risk of cognitive decline conferred by a low nadir CD4 cell count and older age is mixed, with some studies reporting a lack of association (11, 13, 17-21) and others finding and increased risk of decline (16, 22-30). This question remains to be clarified: are older HIV+ individuals with a low nadir CD4 cell count at higher risk of cognitive decline than HIV-negative individuals of the same age? And do methodological differences between existing studies contribute to the discrepancy in the available evidence?

The objectives of this study are threefold: (i) to contribute evidence about the change in cognition over one year among older adults with well-controlled HIV infection and a history of low nadir CD4 cell count, compared to the change in a matched non-HIV sample; (ii) to provide evidence regarding the comparability of different measures of cognitive change used in the neuroHIV literature: and (iii) to contribute evidence to the validity of the classification algorithms by asking whether one of these algorithms applied at baseline provides a better prediction of the magnitude of change over the following year as measured by the NPZ. This was tested for several existing HAND classification algorithms as recent work from our group has found these yield very different rates of impairment (10).

## Methods

### Participants

Between June 2012 and May 2013, 50 clinically stable persons living with HIV were recruited among patients attending the Chronic Viral Illness Service of the McGill University Health Centre in Montreal, Canada. Inclusion criteria were: 40 years or older, stable antiretroviral regimen with an undetectable viral load for at least 6 months, and a nadir CD4 < 200 cells/μL. Exclusion criteria were: history of dementia precluding consent or other neurological condition, current substance use disorder, change in psychoactive medication within the last 2 months, poorly controlled Axis 1 psychiatric disorder and presence of hepatitis C infection, itself associated with cognitive decline (18, 23, 31, 32). Seventeen HIV-negative individuals matched on variables important for neuropsychological (NP) performance, namely sex, age, and education were recruited through advertisements in local gay magazines and community organizations, and in common areas of the hospital.

### Procedure/data collection

The local Research Ethics Board approved the protocol and all subjects provided informed consent. Clinical and socio-demographic information was collected through an interview and a chart review. Participants completed NP testing on two occasions, one year apart. Testing was performed by a trained research assistant, either in English or French, according to a manualized procedure to standardize administration.

### Measurement

We documented NP performance, presence of co-morbidities, and functional status.

### Neuropsychological performance

Following current recommendations, the NP evaluation covered six cognitive domains, with ≥ 2 tests per domain (33). The test battery was developed to support a diagnosis of HAND in Canada, in English and French. Senior neuropsychologists with experience testing in both languages selected the most suitable tests and population norms: Hopkins Verbal Learning Test–Revised (HVLT-R) learning and recall (34); Brief Visuospatial Memory Test-Revised (BVMT-R) learning and recall (35); Tower of London (36); Trail Making Test A and B (TMT-A and B); Stroop (37); Letter/Number Sequencing, Symbol Search, Digit Symbol Coding (38); Spatial Span (39); Letter and Category Fluency (37); Grooved Pegboard dominant and non-dominant hand (40). Alternate forms of the tests were used when appropriate. Intelligence was assessed with the Toni-IV (41).

### Co-morbid conditions

The presence and severity of co-morbid conditions that might preclude a diagnosis of HAND were systematically assessed in those with HIV(42). The Hospital Anxiety and Depression Scale (HADS) was used to screen for the presence of clinically important anxiety and depression(43, 44), with a score ≥ 8 on either scale considered clinically significant, complemented by clinical judgement during testing. Alcohol use was evaluated with the AUDIT-C(45, 46), type and frequency of substances used was assessed by a questionnaire and complemented by a urine toxicology screen. History of CNS opportunistic infection, presence of non-HIV neurological conditions, developmental disability and current systemic disease were documented by chart review. The severity of co-morbid conditions was rated as compatible or contributing according to the guidelines from the 2007 updated research nosology for HAND(33); confounding conditions were excluded. In the HIV-negative group, we documented depression, anxiety and cognitive difficulties.

### Function

Function was assessed with several instruments. In both groups, instrumental activities of daily living (IADL) was assessed with the Older Americans Resources and Services Social Resources Scale (OARS) IADL(47) and the presence of cognitive difficulties in everyday life was documented with the self-reported Perceived Deficit Questionnaire (PDQ), scored 0-80, with a score > 40 indicative of important difficulties (48). In HIV+, adherence was assessed with the Simplified Medication Adherence Questionnaire (SMAQ) and poor adherence was defined as a positive response to any of the qualitative questions (ever forget to take medicine, careless at times about taking your medicine, stop taking medicine when feels worse, failing to take medicine over past week-end), more than two doses missed over the past week, or over 2 days of total non-medication during the past 3 months (49).

### Data analysis

Raw scores on neuropsychological tests were converted to demographically corrected z-scores. Overall NP performance was assessed by the NPZ, calculated by averaging z-scores from all tests, with a lower score indicating worse cognition and a positive value on the change score reflecting an improvement in overall NP performance.

Several algorithms for the diagnosis of HAND in current use in neuro-HIV research were applied. While all algorithms require impairment in ≥ 2 cognitive domains, these methods differ in their definition of domain impairment: Method A: Lowest z-score per domain is > 1 S.D. below norms (50, 51); Method B: Lowest z-score per domain is > 1.5 S.D. below norms (52); Method C: Average z-score for all tests in a specific domain ≥ 1.5 S.D. below norms (53); Method D: one z-score > 1.5 S.D. below norms or 2 tests > 1 S.D. below norms in a domain (54); and Clinical Rating, described elsewhere (55, 56). The classification algorithms were applied to HIV+ and HIV-participants to classify them as cognitively impaired (i.e., compatible with a neurocognitive disorder—presumptively HAND in the case of those with HIV) or not.

Change over one year was defined in two ways: as a change in NPZ or a transition across HAND classification (impaired vs not). We also used Reliable Change Index (RCI) to identify participants who, over the course of one year, made a meaningful change in NPZ value; the RCI is the difference between two assessments divided by a factor related to the standard deviation of each measurement time point and their correlation.

Comparisons were made using 2-sided Fisher’s exact test for variables presented dichotomous, t-test for means of continuous variables deemed normally distributed, with pooled or Satterthwaite variance calculations depending on whether or not the variances were deemed unequal, and Mood’s median test for continuous variables that did not follow a normal distribution.

Distributional parameters were used to characterize the sample and to compare HIV+ and HIV-participants. People were compared on NPZ values at baseline and follow-up by HIV status (HIV+, HIV-) and availability of follow-up data. The proportion of participants classified as impaired (HAND) according to each classification algorithm was calculated, and stability over time was calculated for each participant. As the sample size was small, p-values would not necessarily indicate important differences in proportions of participants impaired. Rather, we used the criterion measure of 10% difference in proportion classified as impaired between HIV+ and HIV-across classification methods and at each time point (57). We used Reliable Change Index (RCI) to identify participants who, over the course of one year, made a meaningful change in NPZ value and tested its association with impairment status according to each classification algorithm at baseline.

## RESULTS

Table 1 shows the personal and clinical characteristics of the sample at the baseline visit.

**Table 1.** Personal and clinical characteristics in HIV-positive and HIV-negative participants at the baseline visit

| Variable | HIV+<br>(n=50) | HIV-<br>(n=17) | p value of<br>difference in means<br>or medians |
| --- | --- | --- | --- |
| **Demographics** | Mean (SD), Median (IQR)<br>or N (%) | Mean (SD), Median (IQR)<br>or N (%) |  |
| Sex, male, N (%) | 46 (92) | 14 (82) | 0.36 |
| Age (years), mean (SD) | 53 (7) | 53 (7) | 0.92 |
| Education (years), mean (SD) | 16 (4) | 16 (4) | 0.90 |
| Ethnicity, Caucasian, N (%) | 38 (76) | 16 (94) | 0.16 |
| Language, French, N (%) | 38 (76) | 11 (65) | 0.36 |
| **Clinical variables** |  |  |  |
| Duration of HIV infection (years), mean (SD) | 16.4 (7.3) |  |  |
| Current CD4+ (cells/μL), median (IQR) | 425.5 (338-648) | | |
| Nadir CD4+ (cells/μL), median (IQR) | 86 (32-143) | | |
| HADS<sup>a</sup> depression score (0-21) |  |  |  |
| median (IQR) | 4 (2-6) | 3 (1-5) | 0.47 |
| depressed ($\geq 8$), N (%) | 7 (14) | 2 (12) | 1.00 |
| HADS<sup>a</sup> anxiety score (0-21) |  |  |  |
| median (IQR) | 6 (5-8) | 6 (4-9) | 0.94 |
| anxious ($\geq 8$), N (%) | 19 (38) | 5 (29) | 0.57 |
| **Overall NP performance** |  |  |  |
| NPZ at baseline, mean (SD) | -0.42 (0.61) | -0.28 (0.97) | 0.59 |
| **Functional impact** |  |  |  |
| Any impairment in IADL, N (%) <sup>d</sup> | 3 (6) | 0 |  |
| Poor medication adherence (SMAQ<sup>b</sup>), N (%) | 33 (66) |  |  |
| PDQ<sup>c</sup> scores (0-80), mean (SD) | 25.4 (13.0) | 21.1 (10.3) | 0.21 |
| PDQ > 40, N (%) | 5 (10) | 0 | 0.32 |

<sup>a</sup> Hospital Anxiety and Depression Scale
<sup>b</sup> Simplified Medication Adherence Questionnaire
<sup>c</sup> Perceived Deficit Questionnaire
<sup>d</sup> Instrumental Activities of Daily Living, Older Americans Resources and Services Social Resources Scale (OARS)

### Comparison of overall cognition, mood and function among HIV+ and HIV-participants at baseline

The HIV+ participants were generally well-educated Caucasian males in their mid-50s, who had been living with HIV for a mean of 16.4 years (range: 2.2 - 26.6 years) at study entry. Current CD4 cell count was in the normal range and nadir CD4 was low at a median of 86 cells/μL, per design. The HIV negative participants were well-matched (3:1) on age, sex, and education. The proportion with clinically important anxiety was high among HIV+ and HIV-groups (38% and 29% respectively); the proportion with depression was lower (HIV+: 14%, HIV-: 12%). At baseline, NPZ scores in HIV+ and HIV-groups were not different. Baseline intelligence was within the normal range in all study participants.

Function at baseline was intact in all HIV-individuals. Among HIV+ participants, one participant reported requiring some help in handling his money, one needing some assistance to do housework and a single participant reported difficulties in two areas, doing housework and preparing meals. Five HIV+ participants endorsed the presence of important cognitive difficulties, including the participant who needed some help to manage finances and the participant who had limitations in both housework and meal preparation. Difficulties with adherence were surprisingly high in this aviremic sample, with 66% reporting some difficulties, mostly with a positive reply to the question “Do you ever forget to take your medicine”. The presence of co-morbid conditions was rated as potentially contributing to cognitive difficulties for two HIV+ individuals with a known attention deficit disorder but no potentially confounding condition was present in any participant.

### Comparison of overall cognition at 1 year according to HIV status and availability of follow-up data

Follow-up at one year was completed by 43/50 HIV+ and 12/17 HIV-participants. Table 2 shows the comparison of the baseline NPZ between those who were retested at 1 year versus those who were not, and between HIV+ and HIV-groups.

**Table 2.** Comparison of the baseline and follow-up NPZ between those with follow-up at 1 year and those without

|  | Mean NPZ (SD) |  |  |  |
| --- | --- | --- | --- | --- |
|  | HIV+ |  | HIV- |  |
|  | N | NPZ, means (SD) | N | NPZ, means (SD) |
|  | NPZ at baseline, comparison of participants with and without follow-up |  |  |  |
| All, baseline NPZ | 50 | -0.42 (0.61) | 17 | -0.28 (0.97) |
| Without follow-up, baseline NPZ | 7 | -0.73 (0.67) | 5 | -0.63 (0.99) |
| With follow-up, baseline NPZ | 43 | -0.37 (0.59) | 12 | -0.14 (0.96) |
| Difference in mean NPZ at baseline between those with and without FU<sup>a</sup> | 50 | 0.36 (0.60)<br>95% CI: -0.13, 0.86<br>p=0.15 | 17 | 0.49 (0.97)<br>95% CI: -0.61, 1.59<br>p=0.36 |

|  | NPZ at 1 year, comparison of HIV+ and HIV- participants |  |  |  |
| --- | --- | --- | --- | --- |
| With follow-up, NPZ at 1 year | 43 | -0.25 (0.52) | 12 | 0.04 (1.00) |
| Mean difference in NPZ at follow-up minus baseline | 43 | 0.12 (0.32)<br>95% CI: 0.02, 0.22<br>p=0.02 | 12 | 0.18 (0.31)<br>95% CI: -0.01, 0.38<br>p=0.06 |
| Difference HIV+ to HIV- at follow-up | -0.06 (0.32), 95% CI: (-0.27, 0.15)<br>p=0.54 |  |  |  |

<sup>a</sup> T-test, p-values from t-tests with pooled variance

Among HIV+ and HIV-, those who were not retested at 1 year were more impaired at baseline than those who were, but the difference was not significant. Change in overall NP performance over one year as measured by the change in NPZ was not different between HIV+ and HIV-participants, with both groups showing a slight improvement in the NPZ at follow-up. The analyses were repeated using the last value carried forward for those missing follow-up; the results remained unchanged (data not shown).

### Comparison of HAND classification and associated change in NPZ, at baseline and 1 year according to HIV status

On NP tests, missing data were minimal except for the Stroop test that could not be completed by seven participants who were color-blind. Table 3 shows the proportion of participants who met impairment criteria for each HAND classification algorithm and evolution over 1 year, according to HIV status. Assigning all missing data as impaired did not change the classification of impairment for any participant.

**Table 3.**
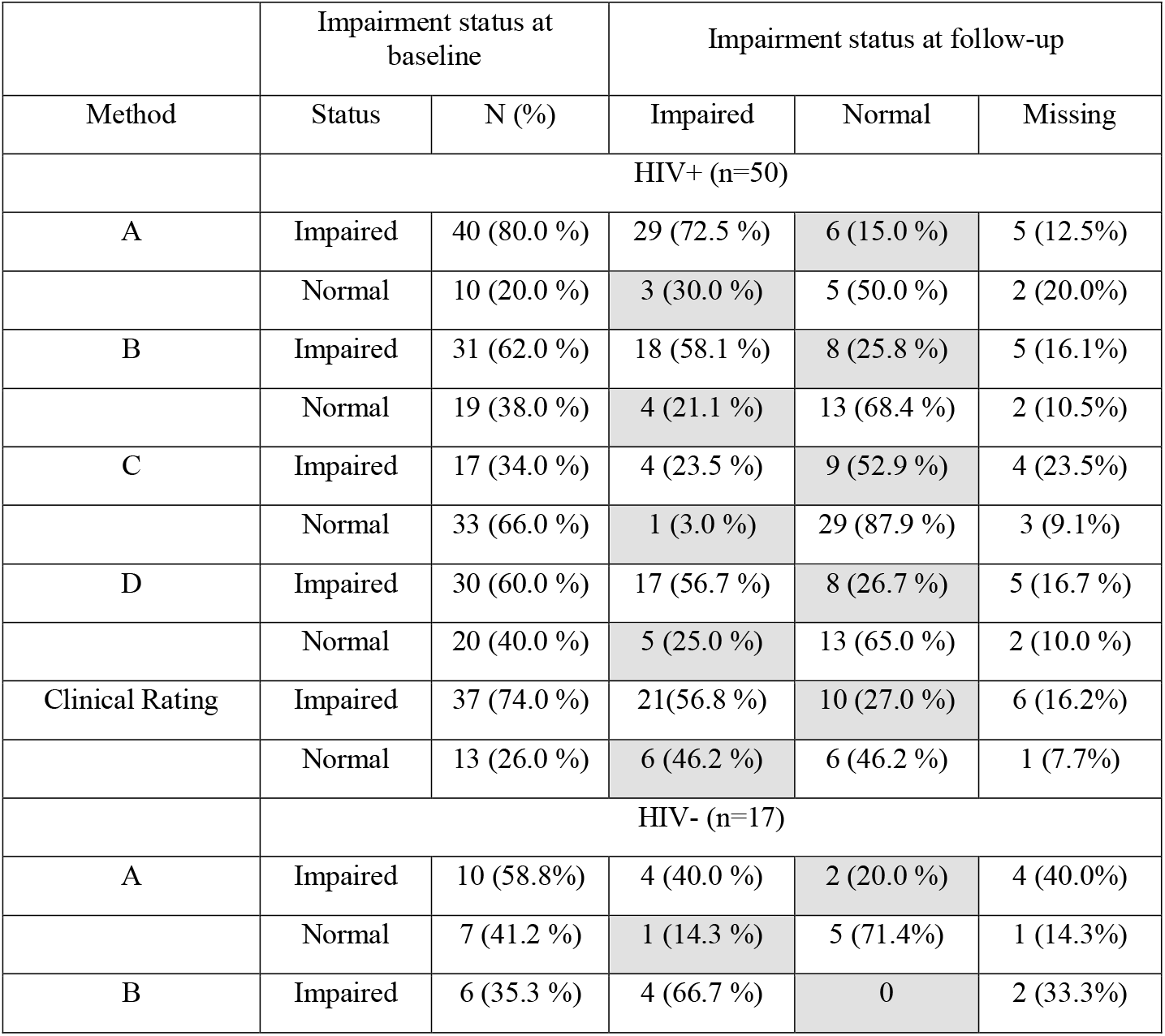
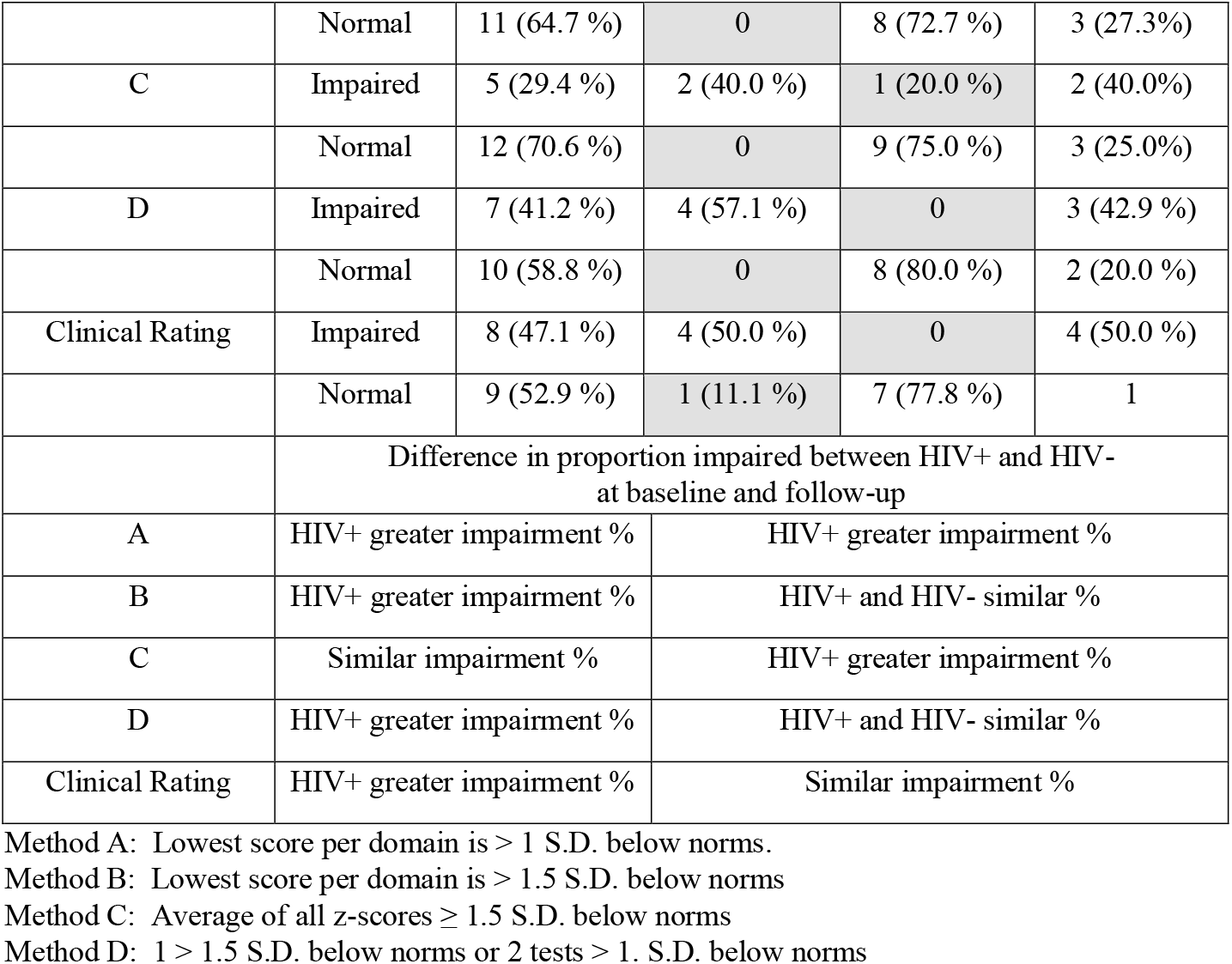
Proportion of participants who met impairment criteria for each HAND classification algorithm and evolution over 1 year, with shaded boxes representing a change in classification between impaired (HAND) versus not.

Among HIV+ individuals, the proportion of impairment at baseline ranged from 34 % (Method C) to a high of 80 % (Method A). The rate of neurocognitive impairment according to these criteria was also high among HIV-participants, ranging from 29.4 % (Method C) to 58.8 % (Method A). Change in impairment status over the follow-up period was quite common and was more often in the direction of improvement, with a transition from impaired to unimpaired. Among the HIV+, the highest stability in impairment classification was seen among those who were classified as normal according to Method C, with 87.9 % remaining normal at follow-up. At the other end of the spectrum, as many as 52.9 % of those classified as impaired at baseline by the same method were classified as normal at follow-up. At baseline, using our criterion measure of a difference of 10%, Methods A, B, D and CR identified people with HIV+ to have a higher prevalence of impairment than people who were HIV-; one year later, only Methods A and C distinguished the two groups. Classification Method A was the only method that consistently distinguished between HIV+ from HIV-over time.

Figure 1 shows the change in NPZ over 1 year for each HAND classification at baseline, with bars in the boxes representing those who were classified as impaired at baseline.

**Figure 1.**
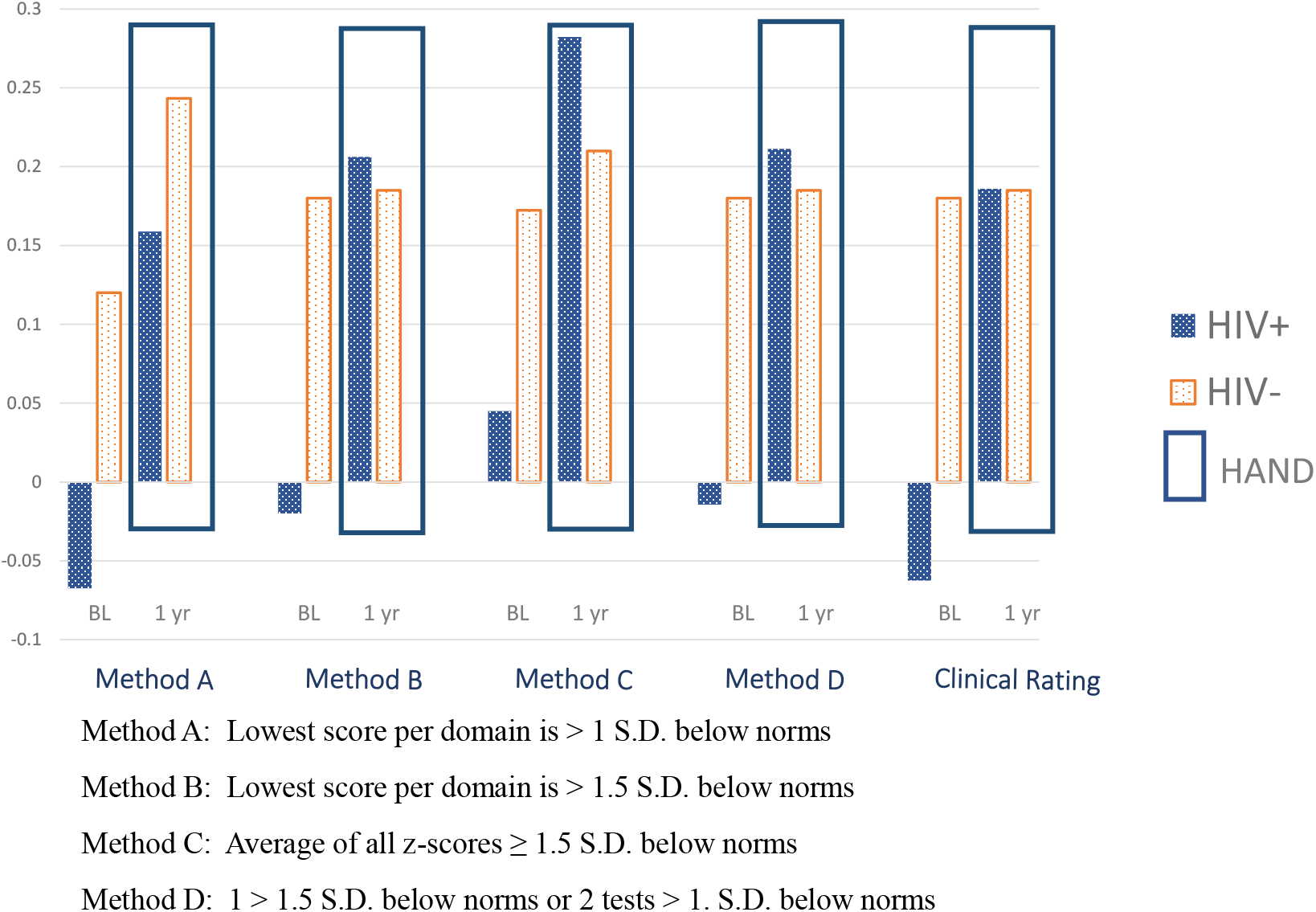
Average change in NPZ according to impairment and HIV status for different classification methods

Declines in NPZ were seen only in HIV+ participants classified as unimpaired at baseline and were greater for those classified as impaired by Methods A and Clinical Rating, but the magnitude of the decline was small, one-tenth of a SD, thus probably not clinically important (58). Improvement in NPZ was seen in several groups, but never exceeded one-third of a SD. No diagnostic algorithm emerged as being clearly superior in predicting the magnitude of cognitive decline at 1 year.

In contrast to the common change in HAND classification, only a single HIV+ participant made a reliable change on the NPZ, so no analysis of association with diagnostic classification was conducted.

## CONCLUSIONS

Among aviremic HIV+ older adults with a history of low nadir CD4 cell count, change in overall NP performance over one year was comparable to that seen among matched HIV-individuals, and a single HIV+ participant made a reliable change on that measure. This is excellent news for survivors of the pre-cART era, adding further evidence that such individuals generally show a trajectory of cognitive change comparable to that of their age- and education-matched HIV-counterparts, at least over the short term.

The application of various existing algorithms for the operationalization of the HAND diagnostic criteria resulted in proportions of impairment at baseline ranging from 34 to 80 % among the HIV+, and from 29.4 to 58.8% among HIV-participants (see Table 3). Four of the five classifications algorithms showed greater prevalence of impairment (> 10% in prevalence) for HIV+ participants compared to HIV-. At follow-up, only classification A distinguished these two groups on prevalence of impairment. Classification A also assigned the greatest proportion of people as impaired at baseline, 80 % for HIV+ and 58.8% for HIV-, raising concerns about the possibility of overdiagnosis.

Change in HAND classification among those with HIV was common in our sample for all classification methods, as was previously seen in studies using a more limited number of methods (16, 59, 60). None of the tested HAND classification algorithms emerged as being superior in identifying those whose cognition will decline over the following year but this is not surprising as, in our sample, improvement in NPZ scores was much more common than decline; decline, when present, was very small and far less than the 0.5 SD usually considered indicative of meaningful change (58). The observed improvement in NPZ scores is likely due to a combination of regression to the mean and practice effects, despite the use of alternate forms of some cognitive tests. The modest changes in NPZ values were often accompanied by a change in HAND classification status. This illustrates the high risk of misclassification associated with use of a binary classification system, especially when individuals are close to the cut-off scores, as seems to have been the case here; this lends support to the recommendation to analyze performance on cognitive tests as a continuous variable in research studies (16, 61). Alternative approaches to identification of cognitive phenotypes of brain involvement are emerging (20) and may shed a new light on the understanding of HIV’s impact on the brain.

Our findings do not completely exclude the possibility that progressive brain dysfunction does take place at a higher rate among selected HIV+ individuals than among matched HIV-individuals. Our measure of overall NP performance, the NPZ, was selected for its sensitivity to a decline of performance in the normal range, in contrast to alternative measures used in neuroHIV research such as the Global Deficit Score (GDS)(62). Despite the selection of this sensitive measure, over a one-year period, we have seen little evidence of neurocognitive decline and a reliable change occurred in a single participant. Brain imaging may provide more sensitive biomarkers for brain changes, although at least some longitudinal data argue that brain atrophy in those with HIV infection on cART does not progress faster than in age-matched controls (63-65), and longer follow-up periods are needed.

Our study adds to the limited evidence regarding the risk for cognitive decline associated with a low nadir CD4 cell count among well-treated HIV+ individuals. A study of middle-aged clinically stable HIV+ individuals (88% aviremic) in Australia found that cognitive decline over an 18-month period, as measured by the Global Change Score approach, did not differ statistically between 96 HIV+ and 44 matched HIV-subjects; while a low nadir CD4 cell count was not an inclusion criteria for their HIV+ participants, the mean value was low, at 181 cells/μL(17). Similarly, in the Multicenter AIDS Cohort Study (MACS), 77% of gay/bisexual male participants on cART with virologic suppression followed over 4 years did not show a progression across HAND categories, and a low nadir CD4 cell count was not associated with an increased risk for HAND progression.

In this study, we classified individuals as impaired (HAND) or not, and did not make a distinction between asymptomatic neurocognitive impairment (ANI) and mild neurocognitive disorder (MND), even though information about function was available. The distinction between ANI and MND hinges on the degree of impairment in activities of daily living but the methods used to assess function vary across research groups and can result in important disparities in those classified as ANI and MND (66). Here, to avoid this methodological problem, the information about function was used to characterize the sample and those with ANI and MND were combined in a category of ‘impaired’. The rate of self-reported functional impairment in the HIV+ group was low except with regards to self-reported adherence. This finding is surprising as all participants were aviremic; however, the importance of these reported difficulties should not be underestimated as good virological control at study entry does not exclude the possibility of viral blips that can have a detrimental effect on disease progression (67).

Our study contributes important information about the impact of the operationalization of cognitive change. Here, we confirm our previous finding that algorithms used by various research groups identify very different groups of individuals (10), such that great caution has to be exercised in comparing findings. This methodological aspect likely contributes to inconsistent findings across studies and constitutes an important impediment to the pooling of data that is necessary for the field to move forward.

One of the strengths of our study is the presence of a demographically matched HIV-group. This allows us to determine if the cognitive decline is greater than expected based on age, education, and lifestyle factors. In the current test-and-treat era, among those with access to cART, the majority of HIV+ individuals have an undetectable viral load. Are those people different from their HIV-counterparts? We find that the role of historical HIV clinical markers may be limited, echoing the conclusion of Bonnet et al. that, among participants in the Aquitaine cohort, the presence of cognitive impairment was not related to HIV nor cART-related variables (50).

Our sample, by design, is not representative of all older HIV+ individuals. It represents a group of special clinical interest, individuals who are well treated and aviremic but present a history of low nadir CD4 cell count. Rates of neurocognitive impairment in this study are high, 34-80 %. While bearing in mind the challenges in comparing studies mentioned above, these rates are comparable to those reported elsewhere. Among 1525 participants in the CHARTER cohort, 39% had HAND. In the NEAD cohort (68) that also selected individuals with a CD4 count <200 cells/μL, Sacktor reported a rate of impairment of 76%, defined by overall neuropsychologist’s impression: however, 77% of the participants had a detectable viremia. Wojna reported a prevalence of HIV-associated cognitive impairment of 77.6% in a group of 49 HIV+ Hispanic women in Puerto Rico who had a nadir CD4 cell count of ≤500 cells/μl (69). Of note, the rates of impairment in our HIV-participants were high as well, a finding reported by others that raises important questions about the validity of current HAND definitions (70-72).

This study has limitations. The control sample was small as it was difficult to recruit normal subjects for this type of testing. In addition, the follow-up period of 1 year was short and, as is usual, there was some loss to follow-up over a year. Those who were not retested at 1 year in the HIV+ and HIV-had lower cognition at baseline than those who were retested; although the difference was potentially meaningful, it was not statistically significant with this sample size. This indicates that in longitudinal studies, those who are most impaired are challenging to retain, with important implications for the results.

In conclusion, in a sample of aviremic HIV+ individuals at potentially higher risk of cognitive impairment as a consequence of older age and nadir CD cell count < 200 cells/μL, the prevalence of HAND ranged from 34-80% depending on the criteria used for classification. Functional impairment was very mild and change in NPZ over one year was not different than that of matched HIV-individuals. The finding that cognitive decline is not inevitable, at least over the short term, is encouraging for persons living with HIV who dread the development of progressive cognitive impairment as they age. It argues for encouraging people with HIV to adhere to cART and to recommendations to maintain brain health for the general adult population such as smoking cessation, regular physical activity and optimal control of glucose and lipids.

## Data Availability

All data produced in the present study are available upon reasonable request to the authors

## Acknowledgement

This study was supported by the CIHR Canadian HIV Trials Network (CTNPT 005). We express our gratitude to the study participants.

## Notes

Competing Interests: the authors have no conflict of interest to report.

### Competing Interest Statement

The authors have declared no competing interest.

### Author Declarations

Ethics committee/IRB of MUHC Research Ethics Board gave ethical approval for this work

## References

1. Cysique LA, Brew BJ. Prevalence of non-confounded HIV-associated neurocognitive impairment in the context of plasma HIV RNA suppression. Journal of neurovirology. 2011;17(2):176–83.

2. Heaton RK, Clifford DB, Franklin DR, Jr., Woods SP, Ake C, Vaida F, et al. HIV-associated neurocognitive disorders persist in the era of potent antiretroviral therapy: CHARTER Study. Neurology. 2010;75(23):2087–96.

3. Jia H, Uphold CR, Wu S, Reid K, Findley K, Duncan PW. Health-related quality of life among men with HIV infection: effects of social support, coping, and depression. AIDS patient care and STDs. 2004;18(10):594–603.

4. Letendre SL, McCutchan JA, Childers ME, Woods SP, Lazzaretto D, Heaton RK, et al. Enhancing antiretroviral therapy for human immunodeficiency virus cognitive disorders. Annals of neurology. 2004;56(3):416–23.

5. Power C, Boisse L, Rourke S, Gill MJ. NeuroAIDS: an evolving epidemic. Can J Neurol Sci. 2009;36(3):285–95.

6. Price RW, Yiannoutsos CT, Clifford DB, Zaborski L, Tselis A, Sidtis JJ, et al. Neurological outcomes in late HIV infection: adverse impact of neurological impairment on survival and protective effect of antiviral therapy. AIDS Clinical Trial Group and Neurological AIDS Research Consortium study team. AIDS. 1999;13(13):1677–85.

7. Tozzi V, Balestra P, Galgani S, Murri R, Bellagamba R, Narciso P, et al. Neurocognitive performance and quality of life in patients with HIV infection. AIDS research and human retroviruses. 2003;19(8):643–52.

8. Vivithanaporn P, Heo G, Gamble J, Krentz HB, Hoke A, Gill MJ, et al. Neurologic disease burden in treated HIV/AIDS predicts survival: a population-based study. Neurology. 2010;75(13):1150–8.

9. Ellis RJ, Rosario D, Clifford DB, McArthur JC, Simpson D, Alexander T, et al. Continued high prevalence and adverse clinical impact of human immunodeficiency virus-associated sensory neuropathy in the era of combination antiretroviral therapy: the CHARTER Study. Arch Neurol. 2010;67(5):552–8.

10. Brouillette MJ, Koski L, Forcellino L, Gasparri J, Brew BJ, Fellows LK, et al. Predicting occupational outcomes from neuropsychological test performance in older people with HIV. AIDS. 2021;35(11):1765–74.

11. Sacktor N, Skolasky RL, Seaberg E, Munro C, Becker JT, Martin E, et al. Prevalence of HIV-associated neurocognitive disorders in the Multicenter AIDS Cohort Study. Neurology. 2016;86(4):334–40.

12. Qu Y, Weinstein A, Wang Z, Cheng Y, Kingsley L, Levine A, et al. Legacy effect on neuropsychological function in HIV-infected men on combination antiretroviral therapy. AIDS. 2022;36(1):19–27.

13. Yuen T, Brouillette MJ, Fellows LK, Ellis RJ, Letendre S, Heaton R, et al. Personalized Risk Index for Neurocognitive Decline Among People With Well-Controlled HIV Infection. Journal of acquired immune deficiency syndromes. 2017;76(1):48–54.

14. Evering TH, Applebaum A, La Mar M, Garmon D, Dorfman D, Markowitz M. Rates of non-confounded HIV-associated neurocognitive disorders in men initiating combination antiretroviral therapy during primary infection. AIDS. 2016;30(2):203–10.

15. Chan P, Kerr SJ, Kroon E, Colby D, Sacdalan C, Hellmuth J, et al. Cognitive trajectories after treatment in acute HIV infection. AIDS. 2021;35(6):883–8.

16. Damas J, Ledergerber B, Nadin I, Tarr PE, Stoeckle M, Kunze U, et al. Neurocognitive course at two-year follow-up in the neurocognitive assessment in the metabolic and aging cohort (NAMACO) study. AIDS. 2021.

17. Gott C, Gates T, Dermody N, Brew BJ, Cysique LA. Cognitive change trajectories in virally suppressed HIV-infected individuals indicate high prevalence of disease activity. PloS one. 2017;12(3):e0171887.

18. Sheppard DP, Woods SP, Bondi MW, Gilbert PE, Massman PJ, Doyle KL, et al. Does older age confer an increased risk of incident neurocognitive disorders among persons living with HIV disease? The Clinical neuropsychologist. 2015;29(5):656–77.

19. Becker JT. Prevalence of cognitive disorders differs as a function of age in HIV virus infection. Aids. 2004;18:11.

20. Paul RH, Cho K, Belden A, Carrico AW, Martin E, Bolzenius J, et al. Cognitive Phenotypes of HIV Defined Using a Novel Data-driven Approach. Journal of Neuroimmune Pharmacology. 2022:1–11.

21. Aung HL, Aghvinian M, Gouse H, Robbins RN, Brew BJ, Mao L, et al. Is There Any Evidence of Premature, Accentuated and Accelerated Aging Effects on Neurocognition in People Living with HIV? A Systematic Review. AIDS and behavior. 2021;25(3):917–60.

22. Ellis RJ, Badiee J, Vaida F, Letendre S, Heaton RK, Clifford D, et al. CD4 nadir is a predictor of HIV neurocognitive impairment in the era of combination antiretroviral therapy. AIDS. 2011;25(14):1747–51.

23. Oliveira NL, Kennedy EH, Tibshirani R, Levine A, Martin E, Munro C, et al. Longitudinal 5-year prediction of cognitive impairment among men with HIV disease. AIDS. 2021;35(6):889–98.

24. Robertson KR, Smurzynski M, Parsons TD, Wu K, Bosch RJ, Wu J, et al. The prevalence and incidence of neurocognitive impairment in the HAART era. AIDS (London, England). 2007;21(14):1915–21.

25. Aung HL, Bloch M, Vincent T, Quan D, Jayewardene A, Liu Z, et al. Cognitive ageing is premature among a community sample of optimally treated people living with HIV. HIV medicine. 2021;22(3):151–64.

26. Aung HL, Gates TM, Mao L, Brew BJ, Rourke SB, Cysique LA. Abnormal cognitive aging in people with HIV: Evidence from Data Integration between two countries’ cohort studies. AIDS (London, England). 2022.

27. Goodkin K, Miller EN, Cox C, Reynolds S, Becker JT, Martin E, et al. Effect of ageing on neurocognitive function by stage of HIV infection: evidence from the Multicenter AIDS Cohort Study. The lancet HIV. 2017;4(9):e411–e22.

28. Maki PM, Rubin LH, Valcour V, Martin E, Crystal H, Young M, et al. Cognitive function in women with HIV: findings from the Women’s Interagency HIV Study. Neurology. 2015;84(3):231–40.

29. Garvey L, Surendrakumar V, Winston A. Low rates of neurocognitive impairment are observed in neuro-asymptomatic HIV-infected subjects on effective antiretroviral therapy. HIV clinical trials. 2011;12(6):333–8.

30. Gawron N, Choiński M, Szymańska-Kotwica B, Pluta A, Sobańska M, Egbert A, et al. Effects of age, HIV, and HIV-associated clinical factors on neuropsychological functioning and brain regional volume in HIV+ patients on effective treatment. Journal of neurovirology. 2019;25(1):9–21.

31. Molsberry SA, Lecci F, Kingsley L, Junker B, Reynolds S, Goodkin K, et al. Mixed membership trajectory models of cognitive impairment in the multicenter AIDS cohort study. AIDS. 2015;29(6):713–21.

32. Grant I, Franklin DR, Jr., Deutsch R, Woods SP, Vaida F, Ellis RJ, et al. Asymptomatic HIV-associated neurocognitive impairment increases risk for symptomatic decline. Neurology. 2014;82(23):2055–62.

33. Antinori A. Updated research nosology for HIV-associated neurocognitive disorders. Neurology. 2007;69(18):1789.

34. Brandt J, Benedict RH. Hopkins verbal learning test--revised: professional manual. Lutz, FL: Psychological Assessment Resources; 2001.

35. Benedict RHB, Schretlen D, Groninger L, Dobraski M Melissa, et al. Revision of the Brief Visuospatial Memory Test: Studies of normal performance, reliability, and validity. Psychological assessment. 1996;8(2):145–53.

36. Culbertson WC, Zillmer EA. Tower of London Drexel University (TOL DX), second edition. Toronto, Canada: Multi-Health Systems; 2005.

37. Delis DC, Kaplan E, Kramer JH. Delis-Kaplan executive function system: The Psychological Corporation; 2001.

38. Wechsler D. Wechsler Adult Intelligence Scale–Fourth Edition: Canadian Technical Manual. Toronto, Ontario: Pearson Canada; 2008.

39. Lafayette Instrument Grooved Pegboard Test: Lafayette Instrument Company; 2002.

40. Heaton R, Miller SW, Taylor MJ, Grant I. Revised comprehensive norms for an expanded Halstead-Reitan Battery: Demographically adjusted neuropsychological norms for African American and Caucasian adults. Lutz, FL: Psychological Assessment Resources. 2004.

41. Brown L, Sherbenou RJ, Johnsen SK. Test of Nonverbal Intelligence, Fourth Edition (TONI-4). Austin, TX2010.

42. Antinori A, Arendt G, Becker JT, Brew BJ, Byrd DA, Cherner M, et al. Updated research nosology for HIV-associated neurocognitive disorders. Neurology. 2007;69(18):1789–99.

43. Zigmond AS, Snaith RP. The hospital anxiety and depression scale. Acta psychiatrica Scandinavica. 1983;67(6):361–70.

44. Olsson I, Mykletun A, Dahl AA. The Hospital Anxiety and Depression Rating Scale: a cross-sectional study of psychometrics and case finding abilities in general practice. BMC Psychiatry. 2005;5:46.

45. Bush K, Kivlahan DR, McDonell MB, Fihn SD, Bradley KA. The AUDIT alcohol consumption questions (AUDIT-C): an effective brief screening test for problem drinking. Ambulatory Care Quality Improvement Project (ACQUIP). Alcohol Use Disorders Identification Test. Archives of internal medicine. 1998;158(16):1789–95.

46. Bradley KA, Bush KR, Epler AJ, Dobie DJ, Davis TM, Sporleder JL, et al. Two brief alcohol-screening tests From the Alcohol Use Disorders Identification Test (AUDIT): validation in a female Veterans Affairs patient population. Archives of internal medicine. 2003;163(7):821–9.

47. Fillenbaum GG. Screening the elderly. A brief instrumental activities of daily living measure. Journal of the American Geriatrics Society. 1985;33(10):698–706.

48. Sullivan JJL, Edgley, K., Dehoux, E. A survey of multiple sclerosis. Part 1: Perceived cognitive problems and compensatory straategy use. Canadian Journal of Rehabilitation. 1990;4:99–105.

49. Knobel H, Alonso J, Casado JL, Collazos J, Gonzalez J, Ruiz I, et al. Validation of a simplified medication adherence questionnaire in a large cohort of HIV-infected patients: the GEEMA Study. AIDS. 2002;16(4):605–13.

50. Bonnet F, Amieva H, Marquant F, Bernard C, Bruyand M, Dauchy FA, et al. Cognitive disorders in HIV-infected patients: are they HIV-related? AIDS. 2013;27(3):391–400.

51. McDonnell J, Haddow L, Daskalopoulou M, Lampe F, Speakman A, Gilson R, et al. Minimal cognitive impairment in UK HIV-positive men who have sex with men: effect of case definitions and comparison with the general population and HIV-negative men. Journal of acquired immune deficiency syndromes (1999). 2014;67(2):120–7.

52. Gisslen M, Price RW, Nilsson S. The definition of HIV-associated neurocognitive disorders: are we overestimating the real prevalence? BMC infectious diseases. 2011;11:356.

53. Meyer AC, Boscardin WJ, Kwasa JK, Price RW. Is it time to rethink how neuropsychological tests are used to diagnose mild forms of HIV-associated neurocognitive disorders? Impact of false-positive rates on prevalence and power. Neuroepidemiology. 2013;41(3-4):208–16.

54. Gates TM, Cysique LA. The Chronicity of HIV Infection Should Drive the Research Strategy of NeuroHIV Treatment Studies: A Critical Review. CNS drugs. 2016;30(1):53–69.

55. Heaton RK, Kirson D, Velin RA, Grant I, Group. tH. The utility of clinical ratings for detecting early cognitive change in HIV infection (pp. 188–206). In: Martin IGA, editor. Neuropsychology of HIV infection: Current research and new directions. New York: Oxford University Press; 1994.

56. Woods SP, Rippeth JD, Frol AB, Levy JK, Ryan E, Soukup VM, et al. Interrater reliability of clinical ratings and neurocognitive diagnoses in HIV. Journal of clinical and experimental neuropsychology. 2004;26(6):759–78.

57. Austin PC. Using the standardized difference to compare the prevalence of a binary variable between two groups in observational research. Communications in statistics-simulation and computation. 2009;38(6):1228–34.

58. Norman GR, Sloan JA, Wyrwich KW. Interpretation of changes in health-related quality of life: the remarkable universality of half a standard deviation. Medical care. 2003;41(5):582–92.

59. Tierney SM, Sheppard DP, Kordovski VM, Faytell MP, Avci G, Woods SP. A comparison of the sensitivity, stability, and reliability of three diagnostic schemes for HIV-associated neurocognitive disorders. Journal of neurovirology. 2017;23(3):404–21.

60. Heaton RK, Franklin DR, Jr., Deutsch R, Letendre S, Ellis RJ, Casaletto K, et al. Neurocognitive change in the era of HIV combination antiretroviral therapy: the longitudinal CHARTER study. Clinical infectious diseases : an official publication of the Infectious Diseases Society of America. 2015;60(3):473–80.

61. Nightingale S, Dreyer AJ, Saylor D, Gisslén M, Winston A, Joska JA. Moving on From HAND: Why We Need New Criteria for Cognitive Impairment in Persons Living With Human Immunodeficiency Virus and a Proposed Way Forward. Clinical Infectious Diseases. 2021;73(6):1113–8.

62. Carey CL, Woods SP, Gonzalez R, Conover E, Marcotte TD, Grant I, et al. Predictive validity of global deficit scores in detecting neuropsychological impairment in HIV infection. Journal of clinical and experimental neuropsychology. 2004;26(3):307–19.

63. Sanford R, Strain J, Dadar M, Maranzano J, Bonnet A, Mayo NE, et al. HIV infection and cerebral small vessel disease are independently associated with brain atrophy and cognitive impairment. AIDS. 2019;33(7):1197–205.

64. Sanford R, Ances BM, Meyerhoff DJ, Price RW, Fuchs D, Zetterberg H, et al. Longitudinal Trajectories of Brain Volume and Cortical Thickness in Treated and Untreated Primary Human Immunodeficiency Virus Infection. Clinical infectious diseases : an official publication of the Infectious Diseases Society of America. 2018;67(11):1697–704.

65. Sanford R, Fernandez Cruz AL, Scott SC, Mayo NE, Fellows LK, Ances BM, et al. Regionally Specific Brain Volumetric and Cortical Thickness Changes in HIV-Infected Patients in the HAART Era. Journal of acquired immune deficiency syndromes. 2017;74(5):563–70.

66. Blackstone K, Moore D, Heaton R, Franklin Jr D, Woods S, Clifford D, et al. Diagnosing symptomatic HIV-associated neurocognitive disorders: self-report versus performance-based assessment of everyday functioning. Journal of the International Neuropsychological Society: JINS. 2012;18(1).

67. Sorstedt E, Nilsson S, Blaxhult A, Gisslen M, Flamholc L, Sonnerborg A, et al. Viral blips during suppressive antiretroviral treatment are associated with high baseline HIV-1 RNA levels. BMC infectious diseases. 2016;16:305.

68. Sacktor N, McDermott MP, Marder K, Schifitto G, Selnes OA, McArthur JC, et al. HIV-associated cognitive impairment before and after the advent of combination therapy. Journal of neurovirology. 2002;8(2):136–42.

69. Wojna V, Skolasky RL, Hechavarria R, Mayo R, Selnes O, McArthur JC, et al. Prevalence of human immunodeficiency virus-associated cognitive impairment in a group of Hispanic women at risk for neurological impairment. Journal of neurovirology. 2006;12(5):356–64.

70. De Francesco D, Underwood J, Post FA, Vera JH, Williams I, Boffito M, et al. Defining cognitive impairment in people-living-with-HIV: the POPPY study. BMC infectious diseases. 2016;16(1):617.

71. Su T, Schouten J, Geurtsen GJ, Wit FW, Stolte IG, Prins M, et al. Multivariate normative comparison, a novel method for more reliably detecting cognitive impairment in HIV infection. AIDS. 2015;29(5):547–57.

72. Underwood J, De Francesco D, Leech R, Sabin CA, Winston A, Pharmacokinetic, et al. Medicalising normality? Using a simulated dataset to assess the performance of different diagnostic criteria of HIV-associated cognitive impairment. PloS one. 2018;13(4):e0194760.

